# How frequent are acute reactions to COVID-19 vaccination and who is at risk?

**DOI:** 10.1101/2021.10.14.21265010

**Authors:** Nancy A. Dreyer, Matthew W Reynolds, Lisa Albert, Emma Brinkley, Tom Kwon, Christina Mack, Stephen Toovey

## Abstract

**Introduction:** Our objective was to describe and compare self-reported side effects of COVID-19 vaccines in the USA.

**Methods:** A web-based registry enrolled volunteers who received a COVID-19 vaccine between March 19 –July 15, 2021. We collected self-reported short-term side effects, medical consultation, hospitalization, and quality of life impact following completed vaccination regimens (Pfizer, Moderna, J&J).

**Results:** We recruited 6,966 volunteers who completed their full course of vaccination (median age 48 years, IQR 35.0-62.0; 83.6% female): Pfizer 3,486; Moderna 2,857; J&J 623. Few (3.1%) sought medical care for post-vaccination side effects. Hospitalization (n=17; 0.3%) and severe allergic reactions (n=39; 0.6%) also were rare. Those with autoimmune disease or lung disease were approximately twice as likely to seek medical care (adjusted odds ratio (aOR) 2.01 [95% CI: 1.39;2.92] and 1.70 [95% CI: 1.12;2.58] respectively). 92.4% of participants reported ≥1 side effect (median 3), with injection site reactions (78.9%), fatigue (70.3%), headache (49.0%) reported most frequently. More side effects were reported after the second dose of two-dose vaccines (medians: 1 vs. 2 for Pfizer and 1 vs. 3 for Moderna for first and second doses respectively) versus 3 for J&J’s single-dose vaccine. For the employed, the median number of workdays missed was one. Diabetics and those vaccinated against influenza were substantially less likely to report ≥3 symptoms (aOR 0.68, 95% CI 0.56,0.82 and aOR 0.82, 95% CI 0.73,0.93, respectively.)

**Discussion:** The total side effect burden was, not unexpectedly, greater with two-dose regimens but all three vaccines appear relatively safe. Very few subjects reported side effects serious enough to warrant medical care or reported post-vaccination hospitalization. While these findings do not address possible long-term effects, they do inform on their short-term safety and tolerability and will hopefully provide some reassurance and positively inform the benefit-risk and pharmacoeconomic assessment for all three vaccines.

**Clinicaltrials.gov NCT04368065**

## Introduction

The United States (U.S.) Food and Drug Administration’s (FDA) regulatory pathway for Emergency Use Authorization (EUA) supports expedited review and authorization of vaccines for health emergencies, but never before have so many vaccines been developed and so quickly. Despite the pandemic proportions of coronavirus disease 2019 (COVID-19) and its heavy toll on society, vaccine hesitancy persists, in part due to concern regarding the “speed and safety” of the vaccines developed to fight COVID-19.[1] Additionally, there is little information about how most people react to COVID-19 vaccinations outside of clinical trials, aside from a constant stream of anecdotal evidence. Reliable population-based scientific evidence may help address concerns by providing granularity on the number, type and severity of side effects and the clinical characteristics of those most impacted.

The objective of this work is to describe and compare self-reported side effects from the three COVID-19 vaccines authorized for use in the U.S. between March 19 and July 15, 2021: two two-dose mRNA vaccines, Pfizer, Inc. and BioNTech (Pfizer) and Moderna, Inc. (Moderna), and a single-dose viral vector vaccine, Janssen Pharmaceuticals Companies of Johnson & Johnson (J&J). [2–4] In addition, we characterize the side effects in terms of their impact on work and life and identify medical and other health-related practices that affect the risk of an acute side effect, such as having been vaccinated for influenza.

## Materials and methods

We used a web-based registry the COVID-19 Active Registry Experience (CARE) that was first launched on April 2, 2020 to study COVID-19 symptoms and severity outside of the hospital setting with the goal of understanding what factors, if any, could mitigate the impact of infection with COVID-19 (www.helpstopCOVID19.com).[5] The protocol and web technology have been updated several times, with the most recent major update in March 2021, updated to record self-reported COVID-19 vaccination and subsequent side effects.

Here we examined CARE participants who reported having received a complete course of COVID-19 vaccination between March 19 –July 15, 2021, either both doses of the Pfizer or Moderna vaccines or a single dose of the J&J vaccine. Participants were recruited via social media targeting adult U.S. residents who received a COVID-19 vaccine. Volunteers provided on-line informed consent. The enrollment and follow-up CARE surveys request information on demographics, medical history, COVID-19-like symptoms [6] and severity, COVID-19 test results, COVID-19 vaccination, and use of prescription and non-prescription medications and dietary supplements. Those who reported having been vaccinated are also asked to provide the date of vaccination for first and second doses, as appropriate, manufacturer, and lot number, if known; patients were instructed to refer to their vaccination card to enhance data accuracy. Respondents are asked about side effects after each vaccination using a drop-down menu of choices along with a free text field where additional information can be provided. We also asked whether the COVID-19 vaccination affected their ability to perform activities of daily living such as bathing or dressing, whether they worked less or missed work due to the vaccine, and if they sought health care or were hospitalized due to these side effects. Missing data were not imputed.

We report individual side effects and the total number of side effects by manufacturer and first or booster shots, as appropriate. For the two-dose vaccines, we also counted all side effects reported at either the first and/or second dose. Side effects that were not listed on the survey but were entered as free text were reviewed. The most frequent free text entries that used related terms (e.g. extreme chills, chills, severe chill) were collapsed and categorized into a new side effect (e.g. chills); we included these in the summation of total side effects. We used logistic regression to estimate the odds of self-reported vaccine side effects by manufacturer. Due to small sample size in some variable categories, we combined categories for the logistic regression model. Multivariate models were adjusted by: age (categorical), gender (female vs. nonfemale (including male, transgender, not disclosed, other)), education (high school or less, some college, 4-year college degree, >4 year college degree), race (White, Black or African American, Other), ethnicity of Hispanic or Latino (yes, no), BMI (underweight or normal weight, overweight, obese, severe obesity), smoker (yes, no), medical conditions including anxiety, autoimmune disease, blood disorder, cardiovascular disease, depression, diabetes, hypertension, insomnia or trouble sleeping, kidney disease, lung disease, and influenza vaccination (yes, no).

## Results

### Participant Characteristics

COVID-19 vaccination was reported by 6,966 volunteers, including 3,486 who received both doses of the Pfizer vaccine, 2,857 who received both doses of the Moderna vaccine, and 623 who received the single-dose J&J vaccine (Table 1). Most CARE participants described themselves as White (90.8%), with additional representation from Asians (1.6%), Black or African American (1.3%) and a small group of people who reported a combination of races (3.6%). Approximately 80% of participants were female, a balance that was consistent across all vaccine manufacturers (range: 80.9-84.1%). A substantial proportion of COVID-19 vaccine recipients reported having anxiety (39.3%), depression (32.4%) and/or insomnia or trouble sleeping (30.7%), and only a small proportion of participants reported being treated for cancer (1.1%), a balance that was generally consistent for these conditions across all vaccines.

**Table 1:** Characteristics at enrollment survey among participants who have completed all regimen doses.

|  | All Participants<br>N=6966 | Pfizer<br>N=3486 | Moderna<br>N=2857 | J&J<br>N=623 |
| --- | --- | --- | --- | --- |
| Age in years (N) | 6966 | 3486 | 2857 | 623 |
| Mean (SD) | 48.3 (15.64) | 47.0 (15.62) | 50.1 (15.69) | 47.1 (14.78) |
| Median [IQR] | 48 [35.0,62.0] | 45 [34.0,61.0] | 51 [36.0,64.0] | 48 [35.0,60.0] |
| Age Group (years) | 6966 | 3486 | 2857 | 623 |
| 18-29 | 781 (11.2) | 433 (12.4) | 260 (9.1) | 88 (14.1) |
| 30-39 | 1768 (25.4) | 966 (27.7) | 678 (23.7) | 124 (19.9) |
| 40-49 | 1067 (15.3) | 551 (15.8) | 402 (14.1) | 114 (18.3) |
| 50-59 | 1241 (17.8) | 581 (16.7) | 519 (18.2) | 141 (22.6) |
| >=60 | 2109 (30.3) | 955 (27.4) | 998 (34.9) | 156 (25.0) |
| Gender | 6966 | 3486 | 2857 | 623 |
| Female | 5821 (83.6) | 2913 (83.6) | 2404 (84.1) | 504 (80.9) |
| Male | 971 (13.9) | 483 (13.9) | 392 (13.7) | 96 (15.4) |
| Transgender | 72 (1.0) | 33 (0.9) | 27 (0.9) | 12 (1.9) |
| Other | 82 (1.2) | 47 (1.3) | 27 (0.9) | 8 (1.3) |
| Not Disclosed | 20 (0.3) | 10 (0.3) | 7 (0.2) | 3 (0.5) |
| Pregnant | 5821 | 2913 | 2404 | 504 |
| Yes | 1121 (19.3) | 662 (22.7) | 398 (16.6) | 61 (12.1) |
| Education | 6949 | 3477 | 2850 | 622 |
| High school or less | 624 (9.0) | 296 (8.5) | 248 (8.7) | 80 (12.9) |
| Some college | 2056 (29.6) | 973 (28.0) | 855 (30.0) | 228 (36.7) |
| 4-yr college degree | 1813 (26.1) | 959 (27.6) | 701 (24.6) | 153 (24.6) |
| >4yr college degree | 2456 (35.3) | 1249 (35.9) | 1046 (36.7) | 161 (25.9) |
| Race | 6957 | 3481 | 2856 | 620 |
| Black or African American | 88 (1.3) | 52 (1.5) | 31 (1.1) | 5 (0.8) |
| White | 6320 (90.8) | 3140 (90.2) | 2614 (91.5) | 566 (91.3) |
| Other** | 549 (7.9) | 289 (8.3) | 211 (7.4) | 49 (7.9) |
| Ethnicity | 6949 | 3474 | 2853 | 622 |
| Hispanic or Latino | 392 (5.6) | 203 (5.8) | 157 (5.5) | 32 (5.1) |
| BMI | 6626 | 3319 | 2719 | 588 |
| Underweight or Normal weight (15.0 - <25.0) | 1778 (26.8) | 922 (27.8) | 703 (25.9) | 153 (26.0) |
| Overweight (25.0- <30.0) | 1857 (28.0) | 913 (27.5) | 787 (28.9) | 157 (26.7) |
| Obese (30.0 - ≤ 40.0) | 2215 (33.4) | 1113 (33.5) | 913 (33.6) | 189 (32.1) |
| Severe Obesity (> 40.0) | 776 (11.7) | 371 (11.2) | 316 (11.6) | 89 (15.1) |
| Smoker | 6484 | 3256 | 2647 | 581 |
| Yes | 611 (9.4) | 275 (8.4) | 257 (9.7) | 79 (13.6) |
| Vaccinated for Influenza | 6677 | 3343 | 2738 | 596 |
| Yes | 4974 (74.5) | 2514 (75.2) | 2111 (77.1) | 349 (58.6) |
| Medical Conditions | 6755 | 3383 | 2768 | 604 |
| Anxiety | 2656 (39.3) | 1348 (39.8) | 1062 (38.4) | 246 (40.7) |
| Autoimmune disease | 843 (12.5) | 406 (12.0) | 359 (13.0) | 78 (12.9) |
| Blood disorder | 173 (2.6) | 87 (2.6) | 75 (2.7) | 11 (1.8) |
| Cardiovascular disease | 407 (6.0) | 175 (5.2) | 186 (6.7) | 46 (7.6) |
| Depression | 2190 (32.4) | 1103 (32.6) | 891 (32.2) | 196 (32.5) |
| Diabetes | 678 (10.0) | 319 (9.4) | 304 (11.0) | 55 (9.1) |
| Hypertension | 1583 (23.4) | 733 (21.7) | 715 (25.8) | 135 (22.4) |
| Insomnia or trouble sleeping | 2074 (30.7) | 1039 (30.7) | 853 (30.8) | 182 (30.1) |
| Kidney disease | 233 (3.4) | 113 (3.3) | 96 (3.5) | 24 (4.0) |
| Lung disease | 671 (9.9) | 307 (9.1) | 301 (10.9) | 63 (10.4) |
| Organ transplant | 18 (0.3) | 12 (0.4) | 6 (0.2) | 0 |
| On Treatment for Cancer? | 6745 | 3377 | 2764 | 604 |
| Yes | 76 (1.1) | 44 (1.3) | 25 (0.9) | 7 (1.2) |

\* The race category of other includes: 24 American Indians, 112 Asians, 4 Hawaiian or Pacific Islander, 249 who selected multiple races and 160 who selected 'other'.

There were small differences in age across vaccine manufacturers (Pfizer recipients were slightly younger, median age of 45 years old compared to 51 years old for Moderna and 48 years old for J&J). Participants vaccinated with J&J included a higher percentage of smokers (13.6%) than those vaccinated with Moderna (9.7%) or Pfizer (8.4%). Participants who received Pfizer or Moderna vaccines had a higher education level, with many having completed a 4-year college degree and most having completed additional education after the college degree (63.5% and 61.3% respectively), compared to recipients of the J&J vaccine (50.5%). Other demographics and medical characteristics appeared to be comparable across the three vaccine manufacturers.

### Vaccine Side Effects

Most COVID-19 vaccine recipients (92.4%) reported having at least one side effect at the first or second dose (80.9% to 95.4% across manufacturers), with a median of 3 side effects, ranging from 3-4 across vaccine manufacturers (Table 2). However, only 3.1% reported seeking medical care for their post vaccination side effects (4.3% for J&J vaccine recipients followed by Moderna (3.3%) and Pfizer (2.8%)). Very few participants reported being hospitalized after vaccination (n=17; 0.3% overall) or had severe allergic reactions (0.6%). Severe allergic reactions differed slightly by vaccine, with more reported for recipients of J&J (1.3%) compared to 0.6% for Moderna and 0.4% for Pfizer.

**Table 2:** COVID-19 vaccine side effects by manufacturer.

|  | All Participants<br>N=6966 | Pfizer<br>N=3486 | Moderna<br>N=2857 | J&J<br>N=623 |
| --- | --- | --- | --- | --- |
| Side Effects | 6966 | 3486 | 2857 | 623 |
| Fever | 2084 (29.9) | 770 (22.1) | 1119 (39.2) | 195 (31.3) |
| Swollen lymph nodes | 788 (11.3) | 392 (11.2) | 353 (12.4) | 43 (6.9) |
| Diarrhea | 69 (1.0) | 37 (1.1) | 27 (0.9) | 5 (0.8) |
| Nausea or vomiting | 1099 (15.8) | 466 (13.4) | 525 (18.4) | 108 (17.3) |
| Chills | 285 (4.1) | 101 (2.9) | 150 (5.3) | 34 (5.5) |
| Injection site reactions | 5493 (78.9) | 2695 (77.3) | 2468 (86.4) | 330 (53.0) |
| Severe allergic reactions | 39 (0.6) | 14 (0.4) | 17 (0.6) | 8 (1.3) |
| Fatigue | 4897 (70.3) | 2312 (66.3) | 2222 (77.8) | 363 (58.3) |
| New or worsening joint pain | 1505 (21.6) | 614 (17.6) | 767 (26.8) | 124 (19.9) |
| New or worsening muscle pain | 2113 (30.3) | 894 (25.6) | 1043 (36.5) | 176 (28.3) |
| Headache | 3416 (49.0) | 1538 (44.1) | 1595 (55.8) | 283 (45.4) |
| Dizziness | 81 (1.2) | 39 (1.1) | 35 (1.2) | 7 (1.1) |
| No. of Side Effects (N) | 6966 | 3486 | 2857 | 623 |
| Mean (SD) | 3.1 (2.00) | 2.8 (1.91) | 3.6 (2.00) | 2.7 (2.09) |
| Median [IQR] | 3 [2.0,5.0] | 3 [1.0,4.0] | 4 [2.0,5.0] | 3 [1.0,4.0] |
| Min - Max | (0 - 9) | (0 - 9) | (0 - 9) | (0 - 8) |
| 0 side-effects | 529 (7.6) | 279 (8.0) | 131 (4.6) | 119 (19.1) |
| 1 to 2 side-effects | 2419 (34.7) | 1443 (41.4) | 789 (27.6) | 187 (30.0) |
| 3 or more side-effects | 4018 (57.7) | 1764 (50.6) | 1937 (67.8) | 317 (50.9) |
| Sought medical care for any side effects | 6473 | 3222 | 2737 | 514 |
| Yes | 201 (3.1) | 90 (2.8) | 89 (3.3) | 22 (4.3) |
| Hospitalized post-vaccination | 6473 | 3222 | 2737 | 514 |
| Yes | 17 (0.3) | 10 (0.3) | 4 (0.1) | 3 (0.6) |
| Trouble with self-care due to side effects (Yes/No) | 6473 | 3222 | 2737 | 514 |
| Yes | 2030 (31.4) | 853 (26.5) | 1037 (37.9) | 140 (27.2) |
| Trouble with self-care due to side effects (Categorical) | 6473 | 3222 | 2737 | 514 |
| Not at all | 4443 (68.6) | 2369 (73.5) | 1700 (62.1) | 374 (72.8) |
| A little | 1166 (18.0) | 509 (15.8) | 562 (20.5) | 95 (18.5) |
| Somewhat | 478 (7.4) | 200 (6.2) | 251 (9.2) | 27 (5.3) |
| A lot | 386 (6.0) | 144 (4.5) | 224 (8.2) | 18 (3.5) |
| Worked less due to side effects | 4648 | 2380 | 1910 | 358 |
| Yes | 1646 (35.4) | 721 (30.3) | 813 (42.6) | 112 (31.3) |

Note: For two-dose vaccine series, data include side effects reported after either first or booster dose. For those who report missing work due to side effects, it is unknown whether time off from work was sanctioned by their employer to facilitate getting vaccinated.

The most frequently reported side effects were injection site reactions (78.9%) and fatigue (70.3%) followed by headache (49.0%). Injection site reactions were reported most often for Moderna vaccine recipients (86.4%), followed by Pfizer (77.3%) and J&J (53.0%). Similarly, fatigue was also reported more frequently by recipients of Moderna (77.8%) than Pfizer (66.3%) or J&J (58.3%) as was headache (55.8% for Moderna compared to 45.4% for J&J and 44.1% for Pfizer. Other reactions included muscle pain (30.3%), fever (29.9%) and joint pain (21.6%) among all participants. Some differences were noted in the reporting of muscle pain and fever, with these events reported most frequently by those who received a Moderna vaccine (36.5% and 39.2%, respectively), followed by J&J (28.3%, 31.3%) and Pfizer (25.6%, 22.1%). Joint pain was also more frequently reported by Moderna vaccine recipients (26.8%) compared to J&J (19.9%) and Pfizer (17.6%).

Those who received a two-dose regimen (mRNA vaccines) reported more side effects at the second dose (Pfizer: 2 and Moderna: 3) than the first (Pfizer: 1 and Moderna: 1), with the median of 3 side effects for the single-dose J&J vaccine more like the second doses of the mRNA vaccines than the first doses of those vaccines (Table 3). Similarly, difficulty with self-care was reported more frequently after the second dose than the first dose for the mRNA vaccines (36.0% and 25.8% for the second doses of Moderna and Pfizer, respectively) than their first doses (17.0% and 11.7% for Moderna and Pfizer, respectively). Of those receiving the J&J vaccine, 27.2% reported trouble with self-care after the single dose. Overall few people sought medical care (3.1%) or were hospitalized (0.3%) after vaccination, with negligible differences between the first and second dose.

**Table 3:** COVID-19 Vaccine side effects, self-care and effect on work, by first or second injection, by first or second injection.

|  | First dose |  | Second Dose |  | J&J<br>N=623 |
| --- | --- | --- | --- | --- | --- |
|  | Pfizer<br>N=3486 | Moderna<br>N=2857 | Pfizer<br>N=3486 | Moderna<br>N=2857 |  |
| Side Effects | 3486 | 2857 | 3486 | 2857 | 623 |
| Fever | 211 (6.1) | 280 (9.8) | 688 (19.7) | 1027 (35.9) | 195 (31.3) |
| Swollen lymph nodes | 160 (4.6) | 160 (5.6) | 332 (9.5) | 286 (10.0) | 43 (6.9) |
| Diarrhea | 17 (0.5) | 5 (0.2) | 20 (0.6) | 22 (0.8) | 5 (0.8) |
| Nausea or vomiting | 158 (4.5) | 167 (5.8) | 369 (10.6) | 449 (15.7) | 108 (17.3) |
| Chills | 20 (0.6) | 16 (0.6) | 81 (2.3) | 134 (4.7) | 34 (5.5) |
| Injection site reactions | 2418 (69.4) | 2245 (78.6) | 2289 (65.7) | 2180 (76.3) | 330 (53.0) |
| Severe allergic reactions | 5 (0.1) | 5 (0.2) | 13 (0.4) | 14 (0.5) | 8 (1.3) |
| Fatigue | 1309 (37.6) | 1194 (41.8) | 2051 (58.8) | 2060 (72.1) | 363 (58.3) |
| New or worsening joint pain | 219 (6.3) | 277 (9.7) | 532 (15.3) | 661 (23.1) | 124 (19.9) |
| New or worsening muscle pain | 333 (9.6) | 376 (13.2) | 756 (21.7) | 912 (31.9) | 176 (28.3) |
| Headache | 788 (22.6) | 751 (26.3) | 1278 (36.7) | 1407 (49.2) | 283 (45.4) |
| Dizziness | 13 (0.4) | 11 (0.4) | 26 (0.7) | 24 (0.8) | 7 (1.1) |
| No. of Side Effects (N) | 3486 | 2857 | 3486 | 2857 | 623 |
| Mean (SD) | 1.6 (1.39) | 1.9 (1.56) | 2.4 (1.92) | 3.2 (2.04) | 2.7 (2.09) |
| Median [IQR] | 1 [1.0,2.0] | 1 [1.0,3.0] | 2 [1.0,4.0] | 3 [2.0,5.0] | 3 [1.0,4.0] |
| Min - Max | (0 - 9) | (0 - 8) | (0 - 9) | (0 - 9) | (0 - 8) |
| 0 side-effects | 633 (18.2) | 350 (12.3) | 538 (15.4) | 240 (8.4) | 119 (19.1) |
| 1 to 2 side-effects | 2120 (60.8) | 1725 (60.4) | 1489 (42.7) | 911 (31.9) | 187 (30.0) |
| More than 3 side-effects | 733 (21.0) | 782 (27.4) | 1459 (41.9) | 1706 (59.7) | 317 (50.9) |
| Sought Medical care for any side effects | 2884 | 2525 | 2964 | 2622 | 514 |
| Yes | 41 (1.4) | 45 (1.8) | 68 (2.3) | 50 (1.9) | 22 (4.3) |
| Hospitalized post-vaccination | 2884 | 2525 | 2964 | 2622 | 514 |
| Yes | 6 (0.2) | 1 (0.0) | 9 (0.3) | 3 (0.1) | 3 (0.6) |
| Trouble with self-care due to side effects (Yes/No) | 2884 | 2525 | 2964 | 2622 | 514 |
| Yes | 337 (11.7) | 430 (17.0) | 764 (25.8) | 943 (36.0) | 140 (27.2) |
| Trouble with self-care due to side effects (Categorical) | 2884 | 2525 | 2964 | 2622 | 514 |
| Not at all | 2547 (88.3) | 2095 (83.0) | 2200 (74.2) | 1679 (64.0) | 374 (72.8) |
| A little | 260 (9.0) | 306 (12.1) | 453 (15.3) | 520 (19.8) | 95 (18.5) |
| Somewhat | 54 (1.9) | 80 (3.2) | 181 (6.1) | 220 (8.4) | 27 (5.3) |
| A lot | 23 (0.8) | 44 (1.7) | 130 (4.4) | 203 (7.7) | 18 (3.5) |
| Missed work or worked less due to side effects | 2141 | 1781 | 2147 | 1764 | 358 |
| Yes | 239 (11.2) | 229 (12.9) | 628 (29.3) | 743 (42.1) | 112 (31.3) |
| Days worked less (n) | 180 | 162 | 443 | 504 | 79 |
| Mean (SD) | 2.3 (3.76) | 2.5 (5.24) | 2.4 (6.48) | 1.8 (2.99) | 2.3 (2.88) |
| Median (IQR) | 1 [1.0,2.0] | 1 [1.0,2.0] | 1 [1.0,2.0] | 1 [1.0,2.0] | 1 [1.0,3.0] |
| Min-Max | (1 - 35) | (1 - 60) | (1 - 90) | (1 - 60) | (1 - 21) |
| 1 day | 105 (3.0) | 82 (2.9) | 277 (7.9) | 328 (11.5) | 43 (6.9) |
| 2 days | 37 (1.1) | 47 (1.6) | 102 (2.9) | 103 (3.6) | 16 (2.6) |
| 3 or more days | 38 (1.1) | 33 (1.2) | 64 (1.8) | 73 (2.6) | 20 (3.2) |
|  | Pfizer<br>N=3486 | Moderna<br>N=2857 | Pfizer<br>N=3486 | Moderna<br>N=2857 |  |
| Days missed work (n) | 165 | 176 | 478 | 592 | 99 |
| Mean (SD) | 1.9 (2.97) | 2.3 (5.25) | 1.9 (5.52) | 1.6 (2.63) | 1.7 (1.92) |
| Median (IQR) | 1 [1.0,2.0] | 1 [1.0,2.0] | 1 [1.0,2.0] | 1 [1.0,2.0] | 1 [1.0,2.0] |
| Min-Max | (1 - 34) | (1 - 60) | (1 - 100) | (1 - 60) | (1 - 16) |
| 1 day | 109 (3.1) | 104 (3.6) | 331 (9.5) | 425 (14.9) | 66 (10.6) |
| 2 days | 34 (1.0) | 48 (1.7) | 101 (2.9) | 117 (4.1) | 21 (3.4) |
| 3 or more days | 22 (0.6) | 24 (0.8) | 46 (1.3) | 50 (1.8) | 12 (1.9) |

Figure 1 shows the odds of specific side effects for each mRNA vaccine (experienced at either first or second dose) compared to the J&J vaccine. After adjustment for potential confounding factors, injection site reactions, swollen lymph nodes, and fatigue were more likely to be reported with the two mRNA vaccines. Specifically, injection site reactions (pain, redness, or swelling at the vaccine injection site) were more likely to be reported by recipients of Pfizer and Moderna compared to J&J (Pfizer: aOR=3.13, 95% CI: 2.58, 3.81; Moderna: aOR=6.60, 95% CI=5.33, 8.18), as were swollen lymph nodes (Pfizer: aOR=1.69, 95% CI=1.20, 2.38; Moderna: aOR=1.97, 95% CI=1.40, 2.77), and fatigue (Moderna: aOR=1.43 95% CI=1.18, 1.74; Pfizer: aOR=2.93 95% CI=2.39, 3.59). Looking at side effects collectively, those who received the Moderna vaccine were more likely to report having three or more side effects (aOR 2.48, 95% CI 2.03, 3.02) compared to the J&J vaccine (Table 4).

**Figure 1:**
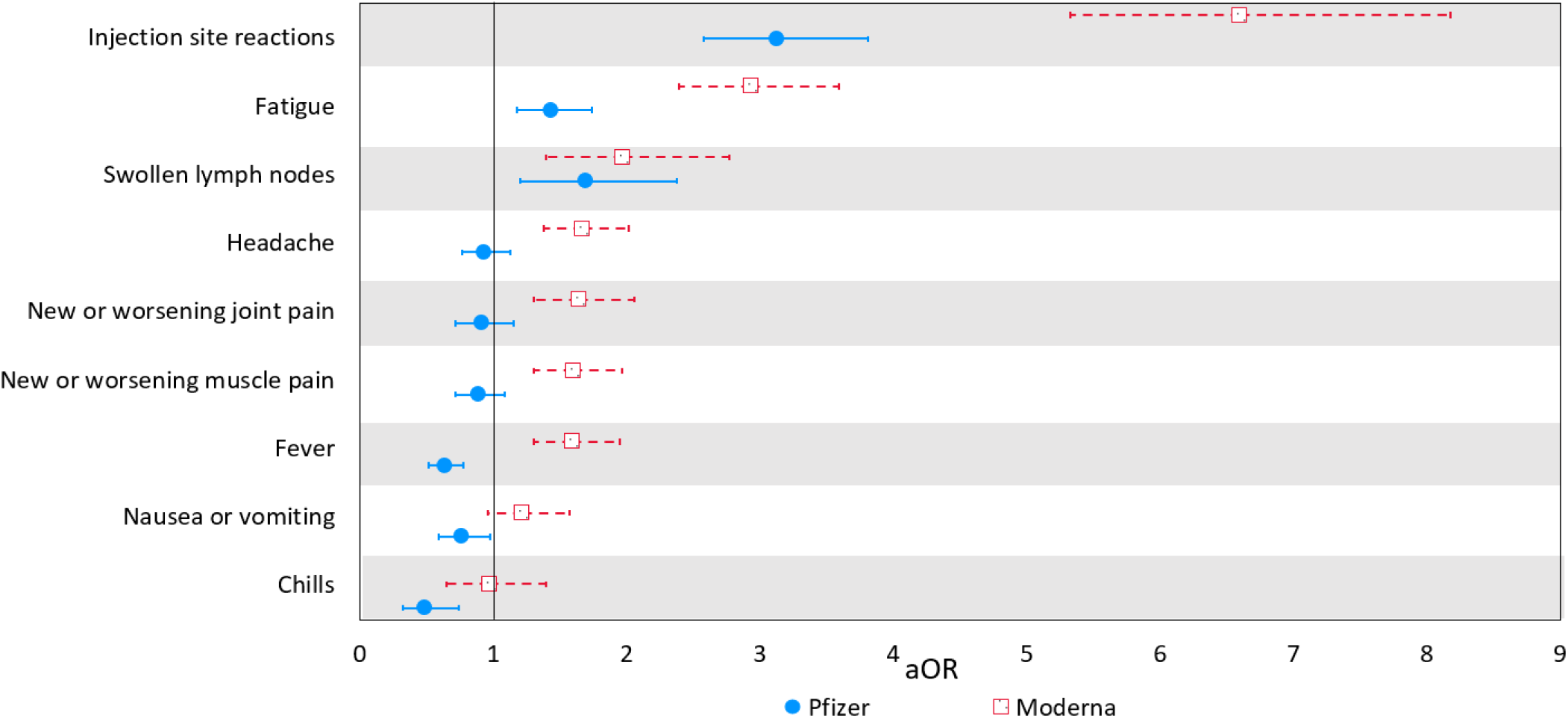
Odds of COVID-19 vaccine side-effects from Moderna and Pfizer vaccines compared to the J&J vaccine (Adjusted Odds Ratios (aOR) and 95% Confidence Intervals (CI)) `Note: For two-dose vaccine series, data include side effects reported after either first or booster dose.` Adjusted ORs are controlled for by age, education, ethnicity, race (White, Black/African American, Other), BMI, receipt of influenza vaccine, gender (female vs. non-female), smoking status, and medical conditions (depression, anxiety, insomnia or trouble sleeping, cardiovascular disease, kidney disease, hypertension, diabetes, and lung disease. Due to the possibility of quasi-complete separation of data points because of small sample sizes in the referent group (J&J) experiencing severe allergic reactions, dizziness, and diarrhea, we do not present results for these outcomes.

**Table 4:**
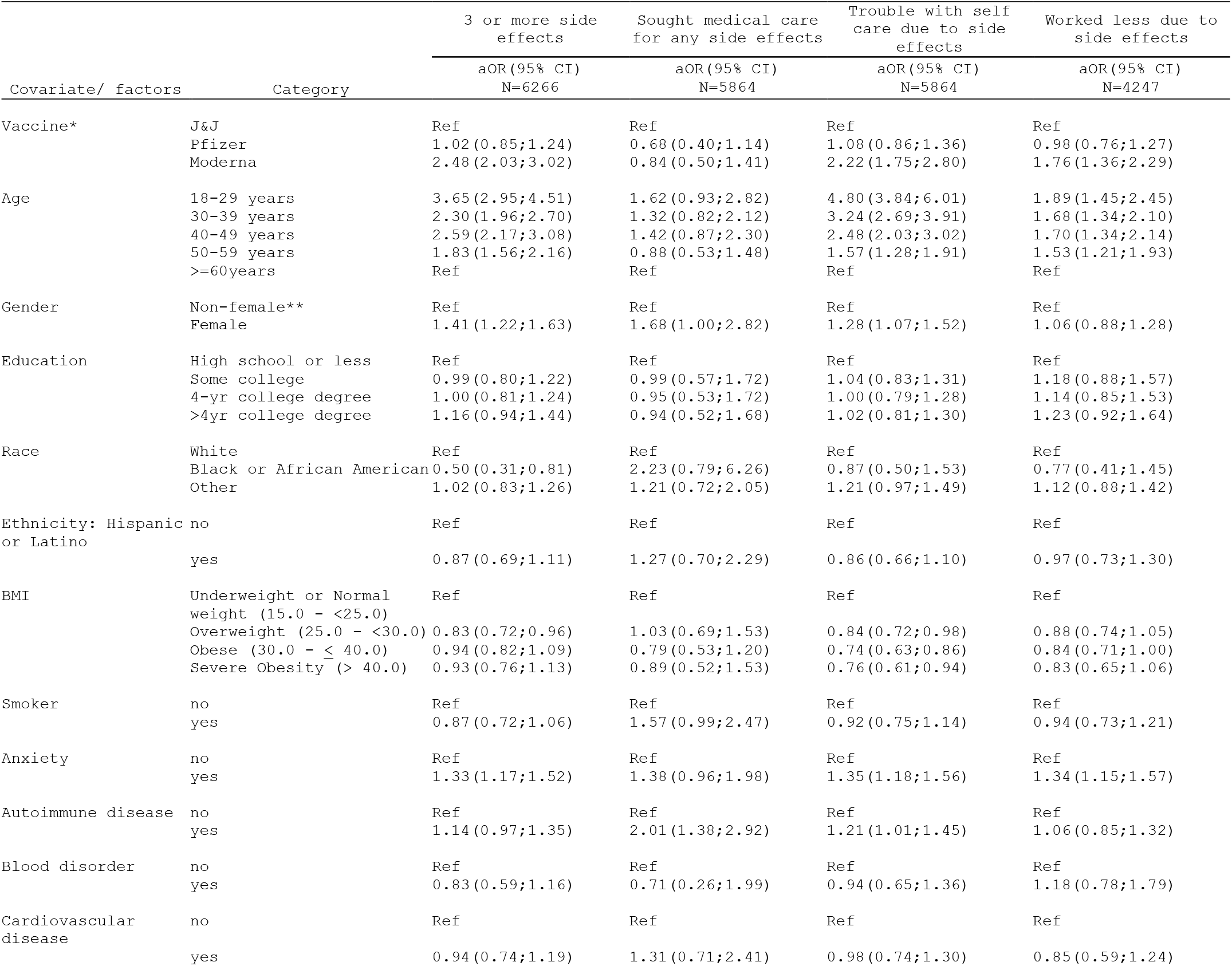
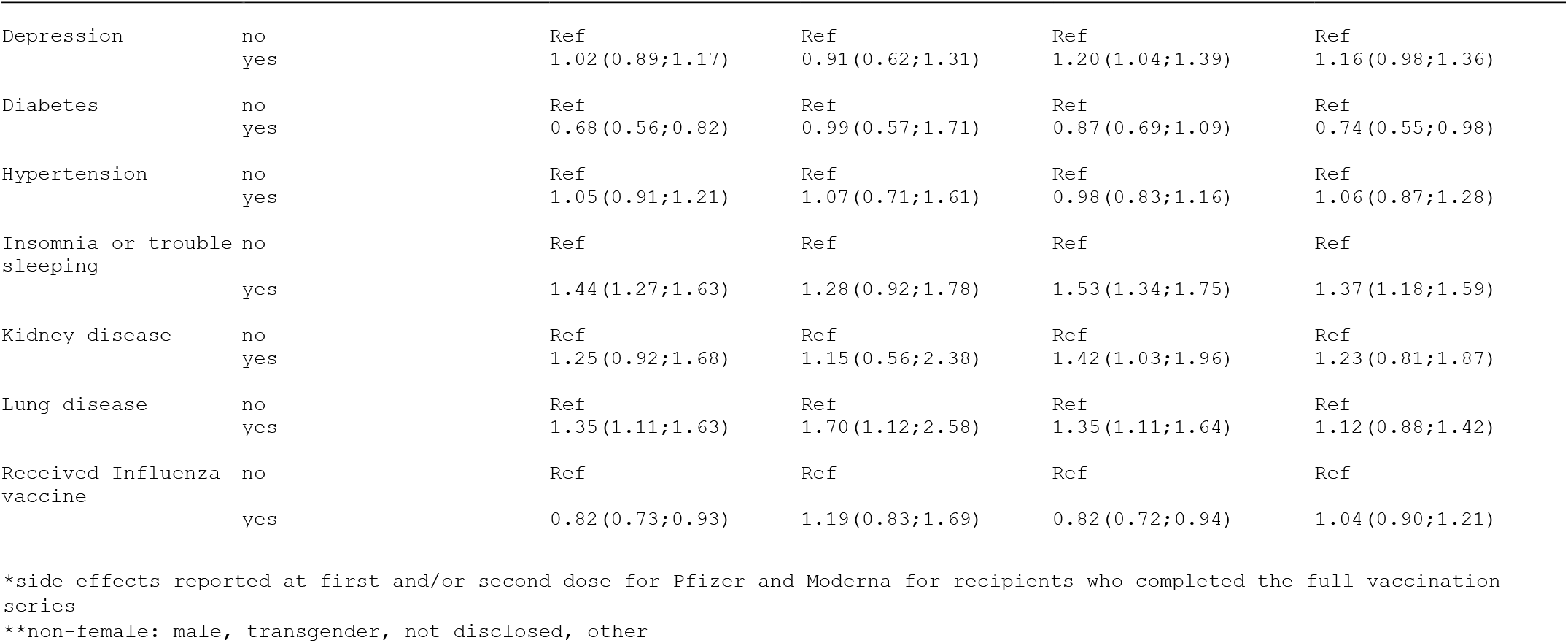
COVID-19 Vaccine side effects by personal characteristics, medical conditions and preventive measures (Adjusted Odds Ratios (aOR) and 95% Confidence Intervals (CI))

Nearly one-third (31.4%) of COVID-19 vaccine recipients reported having trouble taking care of themselves (e.g., with activities such as bathing and dressing) due to vaccination side effects, though only 6.0% described themselves as having had “a lot of trouble” (table 3). The second dose of both mRNA vaccines (Pfizer: 25.8% and Moderna: 36.0%) were associated with greater trouble with self-care than the first doses (Pfizer: 11.7% and Moderna: 17.0%) compared with 27.2% for J&J. Those who received the Moderna vaccine were more than twice as likely to report having trouble caring for themselves (aOR 2.22, 95% CI 1.75, 2.80) compared to the J&J vaccines whereas there was essentially no difference between Pfizer and J&J vaccines (aOR 1.08, 95% CI 0.86, 1.36) (Table 4).

Among those employed 35.4% reported working less after being vaccinated, with a higher proportion of people missing work who received the Moderna vaccine (42.6%) than those who received either the J&J (31.3%) or Pfizer vaccines (30.3%). Of those employed, the median days missed from work was one for all vaccines.

In general, females reported experiencing more side effects and more trouble with self-care than men and others who described themselves as transgender or preferred not to disclose their gender (aOR 1.41 95 CI 1.22, 1.63), though they did not report working any less. Younger people were more likely to report three or more side effects and to report problems caring for themselves than older people, with those ages 18-29 more than three times as likely to report three or more side effects (aOR 3.65, 95% CI 2.95,4.51) and nearly five times as likely to report having trouble with self-care (aOR 4.80, 95% CI 3.84 6.01). Black or African American participants were less likely to report three or more symptoms (aOR 0.50, 95% CI 0.31, 0.81). For people reporting a high BMI (≥30), there was little difference in the proportion of participants reporting three or more symptoms, seeking medical consultation or working less than those who had lower BMI.

For self-reported medical conditions, those with autoimmune disorders were more likely to report trouble with self-care (aOR 1.21, 95% CI 1.01, 1.45) and to have sought medical care more often for side effects (aOR 2.01, 95% CI 1.38, 2.92) than those without autoimmune disorders. Those who reported having insomnia or lung disease also were more likely to report three or more side effects (aOR 1.44, 95% CI 1.27, 1.63 for insomnia, and aOR 1.35, 95% CI 1.11, 1.63 for lung disease) and report having more trouble caring for themselves (aOR 1.53, 95% CI 1.34, 1.75 for insomniacs, and aOR 1.35, 95% CI 1.11, 1.64 for those with lung disease) than those without insomnia or lung disease. In contrast, those with diabetes were less likely to report having three or more symptoms (aOR 0.68, 95% CI 0.56, 0.82) than those without diabetes.

Those who reported having been vaccinated for influenza were less likely to report three or more symptoms (aOR 0.82, 95% CI 0.73, 0.93) or to have trouble caring for themselves (aOR 0.82, 95% CI 0.72, 0.94) than those who did not report having been vaccinated for influenza.

## Discussion

These registry data are consistent with other real-world studies that document few serious side effects from currently authorized COVID-19 vaccines, and also support anecdotal reports describing mostly mild side effects with rapid resolution.[7–9] In this cohort, only 3.1% of vaccine recipients sought medical care for side effects post-vaccination, and almost none (0.3%) were hospitalized. Nonetheless, 57.7% of CARE participants reported experiencing three or more side effects. Roughly one-third (31.4%) of COVID-19 vaccine recipients attributed having some amount of trouble taking care of themselves for the short period immediately after the vaccine due to vaccination side effects and 35.4% of employed participants reported working less, for a single day, due to side effects, noting that the median time lost from work of only one day may reflect employer practices of giving time off from work to encourage vaccination. It seems reasonable to conclude that work time lost from vaccination is far less impactful than work time lost as a consequence of COVID-19 illness.

When comparing the three vaccine manufacturers, Moderna appears to be the most reactogenic overall, with those who received a Moderna vaccine reporting more side effects than those who received Pfizer or J&J vaccines. Second doses of mRNA vaccines were more reactogenic than first doses and these second doses had higher rates of trouble with self-care compared with the first doses of Pfizer and Moderna.

Given the difficulty of comparing two-dose and single-dose regimens to understand side effects, our data enables some degree of comparison between the mRNA and viral vector vaccine classes. When the mean number of side effects per participant is compared, the J&J viral vector vaccine lies somewhere between that observed for the first and second doses of both mRNA vaccines. Thus, per ‘shot’ one might conclude that there is not a great difference in overall tolerability between the two classes, but that the total side effect burden is greater, as might be expected, with a two-dose regimen. This information may be of interest as vaccine supply and choice increase.

An interesting and biologically curious observation was that CARE participants who reported also having had a vaccination for influenza were less likely to report experiencing three or more side effects from COVID-19 vaccination, and had fewer problems taking care of themselves. Although these findings may be associated with health-seeking behavior of influenza vaccine recipients or a greater understanding of common vaccination side effects, there is some evidence that influenza vaccine effectiveness does not vary according to health-seeking behavior,[10] adding credence to the possibility that this may be a true biologic effect.

An increased number of side effects were reported for those with depression, anxiety, insomnia, lung disease, and autoimmune disease. We have previously reported that people with autoimmune disorders appeared to be particularly vulnerable to COVID-19, and their response to COVID-19 vaccination may possibly relate to this susceptibility.[11] In contrast, those with diabetes were less likely than others to report vaccine side effects. The lower reporting rates for diabetics may be influenced by an overlap in potential side effects with their metabolic disorder, or they may have found the vaccine associated side effects rather insignificant as a result. Alternatively, it could be that the progression to severe disease in diabetics has an immunological component that is being reflected in a different immunological reaction to the vaccine, or to the presence of actual virus.

It is important to keep in mind that online registries that survey volunteers without intervention or collaboration from medical care providers rely on the goodwill of participants to contribute information faithfully and accurately, and the results lack clinical or other confirmation. For example, although CARE participants reported their vaccine receipt, including manufacturer and lot number, there was no independent verification of such receipt – a condition that would have been challenging due to the diverse locations used for vaccine administration, the lack of centralized reporting and widespread handwritten vaccine information which would require manual coding. That said, the validity of reports from lay people of medication use has been demonstrated,[12] and as such, reporting quality is likely to apply at least as well to vaccine reporting.

Also, although our findings are derived from community volunteers who responded to internet-based advertisements and posts from medical, patient and other community organizations, this study population has broad geographic representation across the U.S., with participants from all 50 states (with most participants from New York and California, followed by Illinois, Ohio, Pennsylvania, Florida and Texas).

But CARE participants are undoubtedly not representative of the entire US population, including those without internet access nor the free time to participate in surveys. CARE participants have a median age of 48 years, which is consistent with research showing a greater degree of altruism in older than younger adults. More side effects may have been reported by CARE participants than others due to the fact that they self-selected into the registry whereas those with no side effects may be less motivated to enroll in a study. Considering the high proportion of CARE participants who reported suffering from anxiety (39.3%), these reports also may reflect the experience of people who may be more attuned to how they react to vaccination. Nonetheless, comparisons within the CARE population, such as between vaccine manufacturers and by various medical and other personal characteristics, should be broadly generalizable. It is unlikely that there was differential participation according to vaccine manufacturer or that responses selectively exaggerated reactions to one vaccine over another.

Further, although nearly 7,000 people reported their vaccination experience through CARE, the sample is too small to support detection of rare events nor to study rare subgroups. For example, we were not able to separately examine vaccine side effects for transgender people and others who did not wish to report their gender or whose gender did not conform to any of the choices presented in this study. Nonetheless, sensitivity analyses of our statistical models revealed little differences when restricted to those who reported their gender either as male or female.

## Conclusion

On the whole, these community-based insights combined with other information that is accumulating about the effectiveness and safety of COVID-19 vaccines may provide a framework for guiding decision making on vaccine options and addressing vaccine hesitancy, including in particular vulnerable populations, such as diabetics and those with autoimmune disorders. Importantly, these findings provide a patient-centric estimation of vaccine burden by vaccine type and may help inform patients and health care providers on expected side effects and any potential short-term impact on their ability to perform daily activities or work. As CARE registry data continue to accrue, additional analyses may provide insight into real-world effectiveness of COVID-19 vaccines against COVID-19 infection in the context of both presence and severity of break-through infections, as well as more information on how various subgroups of special interest are impacted.

These data may also be useful in the context of current debates about whether COVID-19 vaccination should be made mandatory, at least for certain groups, e.g., healthcare workers and workers in residential care facilities. One of the discussion points in this debate concerns the safety and tolerability of the current COVID-19 vaccines, especially those which have been granted EUA only. While these data currently cannot address the long-term safety and tolerability of these vaccines, vaccines generally display the bulk of their side effects in the very short-term, often a day or less.[13–14] Our findings inform on the short-term safety and tolerability of these vaccines, and will hopefully provide some reassurance on their adverse effects.

## Data Availability

These data are not publicly available but indications of interest for collaboration are welcomed.

## Ethics approval and informed consent

This study was approved by Institutional Review Board and registered at Clinicaltrials.gov NCT04368065 and EU PAS register EUPAS36240. All participants provided informed consent online.

## Consent for publication

Consent for publication of research finding was provided online.

## Data availability

Due to data privacy and security regulations the researchers are not able to share participant level data.

## Acknowledgements

We thank Savitha Pallipuram for leading the tech team, Damir Mohamed for his contributions to the statistical analysis, and Alex Secora for his thoughtful editing contributions.

## References

[1] Bhat, A., Browning-McNee, L. A., Ghauri, K., & Winckler, S. (2021). COVID-19 vaccine confidence project. Journal of the American Pharmacists Association : JAPhA, S1544-3191(21)00273-9. Advance online publication. https://doi.org/10.1016/j.japh.2021.06.006

[2] Centers for Disease Control and Prevention (CDC). Pfizer-BioNTech COVID-19 Vaccine Overview and Safety. https://www.cdc.gov/coronavirus/2019-ncov/vaccines/different-vaccines/Pfizer-BioNTech.html

[3] Centers for Disease Control and Prevention (CDC). Moderna COVID-19 Vaccine Overview and Safety. https://www.cdc.gov/coronavirus/2019-ncov/vaccines/different-vaccines/Moderna.html

[4] Centers for Disease Control and Prevention (CDC). Johnson & Johnson’s Janssen COVID-19 Vaccine Overview and Safety. https://www.cdc.gov/coronavirus/2019-ncov/vaccines/different-vaccines/janssen.html

[5] Dreyer, N. A., Reynolds, M., DeFilippo Mack, C., Brinkley, E., Petruski-Ivleva, N., Hawaldar, K., Toovey, S., & Morris, J. (2020). Self-reported symptoms from exposure to Covid-19 provide support to clinical diagnosis, triage and prognosis: An exploratory analysis. Travel medicine and infectious disease, 38, 101909. https://doi.org/10.1016/j.tmaid.2020.101909.

[6] Symptoms of Coronavirus. Center for Disease Control and Prevention (CDC). Accessed online on January 15, 2021: https://www.cdc.gov/coronavirus/2019-ncov/symptoms-testing/symptoms.html

[7] Gee, J., Marquez, P., Su, J., Calvert, G. M., Liu, R., Myers, T., Nair, N., Martin, S., Clark, T., Markowitz, L., Lindsey, N., Zhang, B., Licata, C., Jazwa, A., Sotir, M., & Shimabukuro, T. (2021). First Month of COVID-19 Vaccine Safety Monitoring - United States, December 14, 2020-January 13, 2021. MMWR. Morbidity and mortality weekly report, 70(8), 283–288. https://doi.org/10.15585/mmwr.mm7008e3

[8] Chapin-Bardales, J., Gee, J., & Myers, T. (2021). Reactogenicity Following Receipt of mRNA-Based COVID-19 Vaccines. JAMA, 325(21), 2201–2202. https://doi.org/10.1001/jama.2021.5374

[9] Shay, D. K., Gee, J., Su, J. R., Myers, T. R., Marquez, P., Liu, R., Zhang, B., Licata, C., Clark, T. A., & Shimabukuro, T. T. (2021). Safety Monitoring of the Janssen (Johnson & Johnson) COVID-19 Vaccine - United States, March-April 2021. MMWR. Morbidity and mortality weekly report, 70(18), 680–684. https://doi.org/10.15585/mmwr.mm7018e2

[10] Jackson, M. L., & Nelson, J. C. (2013). The test-negative design for estimating influenza vaccine effectiveness. Vaccine, 31(17), 2165–2168. https://doi.org/10.1016/j.vaccine.2013.02.053

[11] Dreyer, N., Petruski-Ivleva, N., Albert, L., Mohamed, D., Brinkley, E., Reynolds, M., & Toovey, S. (2021). Identification of a Vulnerable Group for Post-Acute Sequelae of SARS-CoV-2 (PASC): People with Autoimmune Diseases Recover More Slowly from COVID-19. International journal of general medicine, 14, 3941–3949. https://doi.org/10.2147/IJGM.S313486

[12] Dreyer, N. A., Blackburn, S. C., Mt-Isa, S., Richardson, J. L., Thomas, S., Laursen, M., Zetstra-van der Woude, P., Jamry-Dziurla, A., Hliva, V., Bourke, A., & de Jong-van den Berg, L. (2015). Direct-to-Patient Research: Piloting a New Approach to Understanding Drug Safety During Pregnancy. JMIR public health and surveillance, 1(2), e22. https://doi.org/10.2196/publichealth.4939

[13] Lindsey, N. P., Rabe, I. B., Miller, E. R., Fischer, M., & Staples, J. E. (2016). Adverse event reports following yellow fever vaccination, 2007-13. Journal of travel medicine, 23(5), 10.1093/jtm/taw045. https://doi.org/10.1093/jtm/taw045

[14] Lane, J. M., Ruben, F. L., Neff, J. M., & Millar, J. D. (1969). Complications of smallpox vaccination, 1968. The New England journal of medicine, 281(22), 1201–1208. https://doi.org/10.1056/NEJM196911272812201

